# Discussions about Goals of Care in the Emergency Department: a Qualitative Study of Emergency Physicians’ Opinions Using the Normalization Process Theory

**DOI:** 10.1101/2024.07.26.24310500

**Authors:** Fannie Péloquin, Emile Marmen, Véronique Gélinas, Ariane Plaisance, Maude Linteau, Audrey Nolet, Nathalie Germain, Patrick Archambault

**Affiliations:** Department of Family Medicine and Emergency Medicine, Université Laval, Québec, Canada; Centre intégré en santé et en services sociaux de Chaudière-Appalaches, Lévis, Canada; Sciences de la santé, Université du Québec à Rimouski, Lévis, Québec, Canada; Centre de recherche intégrée pour un système apprenant en santé et services sociaux de Chaudière-Appalaches, Lévis, Québec, Canada; VITAM - Centre de recherche en santé durable, Québec, Québec, Canada

**Keywords:** Goals of care discussions (GCD), Advanced care planning, Emergency Medicine, Qualitative research, Normalization Process Theory

## Abstract

**Purpose:** We explored emergency department (ED) physicians’ opinions about the feasibility of leading goals of care discussions (GCD) in their daily practice.

**Method:** This qualitative study was based on the Normalization Process Theory (NPT). We conducted semi-structured interviews between April and May 2018 with a convenience sample of ten emergency physicians from one academic ED (Lévis, Canada) and aimed to reach data saturation. Using a mixed deductive and inductive thematic analysis, two authors codified the interviews under the four NPT constructs: coherence, cognitive participation, collective action, and reflexive monitoring. We calculated a kappa statistic to measure inter-rater agreement.

**Results:** We interviewed 10 emergency physicians. No new ideas emerged after the ninth interview and the inter-rater agreement was substantial. Fourteen themes were identified as factors influencing the feasibility of implementing GCD: (1) interpersonal communication, (2) efficiency of care, (3) anxiety generated by the discussion, (4) identification of an acute deterioration leading to the GCD, (5) meeting of the clinician, patient, and family, (6) importance of knowing the patient’s goals of care before medical handover, (7) lack of training, (8) availability of protocols, (9) heterogeneous prioritization for leading GCD, (10) need to take action before patients consult in the ED, (11) need to develop education programs, (12) need for legislation, (13) need to improve the ED environment and human resources, and (14) selective systematization of GCD for patients.

**Conclusion:** Goals of care discussions are possible and essential with selected ED patients. Physicians identified outstanding needs to normalize GCD in their practice: education for both themselves and patients on the concept of GCD, legislative action for the systematization of GCD for patients, and proactive documentation of patients’ preferences pre-ED. Patient, clinician and system-level policy-making efforts remain necessary to address these needs and ensure the normalization of GCD in emergency physicians’ daily practice as suggested by clinical guidelines.

**Clinician’s capsule:**

1. **What is known about the topic?** Goals of care discussions are important to provide care aligned with patients’ values and medical preferences.
2. **What did this study ask**? According to emergency physicians, are goals of care discussions feasible in the emergency department?
3. **What did this study find?** Goals of care discussions are essential and possible if patient, clinician and system-level policymaking structured efforts are deployed.
4. **Why does this study matter to clinicians**? This study identified action items to improve the implementation and quality of goals of care discussions in the emergency department.

## Introduction

Canada is facing a significant challenge with its aging population, putting pressure on provincial health systems nationwide to respond to an increasing prevalence of age-related illnesses [1]. Unfortunately, increasing numbers of non-beneficial aggressive life-sustaining therapies are also being used near the end of life [2] . Despite this increase in health service use, older adults prioritize their comfort and home-based care near their end-of-life [3]. Experts recommend that acute care professionals improve the frequency and quality of their goals of care discussions (GCD) with older adult patients [4]. GCD allow for health professionals to provide care congruent to patient values and to improve end-of-life care [5]. The emergency department (ED) is a unique environment where time pressures, lack of human resources, overcrowding, and lack of access to important background health information are barriers to establishing a trustful relationship with patients [6–8]. Health organizations across Canada have made recommendations to support and lead earlier and better GCD with older adult patients [9]. In Québec, the *Institut National d’excellence en santé et en services sociaux (INESSS)* has published guidelines recommending emergency physicians to establish goals of care with selected patients [10,11]. In 2016, the guidelines introduced a standardized GCD form featuring four goals of care, and an iterative model to support the implementation of an interprofessional GCD process in all health and social services organizations across the province. A 2024 update to this guideline describes three goals of care but is undergoing pilot testing in select centers and the four-goal model is still widely used [11]. Considering these provincial guidelines, this study aimed to explore emergency physicians’ opinions about leading GCD in their ED with targeted patients.

## Methods

### Study Design and Time period

This study was a cross-sectional qualitative study of ED physicians’ opinions about leading GCD based on the Normalization Process Theory (NPT) [12]. The NPT is a middle-range theory used to explain the sustainability of implementing complex healthcare interventions. We conducted interviews between April and May 2018. This project was approved by the CISSS-CA research ethics board (2018-487) and informed consent was obtained for all participants. We report our results using the Standard for Reporting Qualitative Research guidelines (SRQR) [13] (See Appendix 1.

### Study Setting

The study was conducted in a single academic ED (Hôtel-Dieu de Lévis) in Lévis, Québec, Canada, which is part of the *Centre intégré de santé et de services sociaux de Chaudière-Appalaches (CISSS-CA)*.

### Population and Sample Size

After presenting the project at a departmental meeting, we sent an email invitation to all emergency physicians in the department (n = 31) to participate on a voluntary basis. Those interested contacted the primary author. We established the sample size using an estimation of the number of participants needed to reach theoretical data saturation [14]. We recruited a convenience sample of ten participants, allowing for the recruitment of additional participants if data saturation was not obtained.

### Data Collection and Analysis

Data collection and analysis was led by a six-person team composed of an emergency medicine resident interested in GCD (FP), one medical student with a law degree (EM) and another medical student with palliative care experience (AN), a PhD student with experience in advance care planning and in applying the NPT (AP) [15,16], a research coordinator with experience in qualitative research (VG) [17] and an emergency physician with experience in qualitative research (PA) [18,19]. Interviews were led by FP who had previously collaborated with the participants as a trainee, contributing to a climate of trust and openness among the participants. This also allowed the interviewer to have firsthand experience of the context in which the participants lead GCD.

She conducted semi-structured interviews (40 to 90 minutes) over the phone or in-person with participants based on their preference. All interviews were audio recorded. The interview guide was based on the four constructs of the NPT: coherence, cognitive participation, collective action, and reflexive monitoring [12] (See Appendix 2). Coherence refers to the sense-making done individually and collectively when faced with the operationalization of a new practice. Cognitive participation is the relational work done to build and sustain a community of practice around a new complex intervention. Collective action is the operational work done to enact a complex healthcare intervention. Reflexive monitoring is appraisal work that is done to assess and understand the ways that a new set of practices affect oneself and others [12].

The primary author (FP) and two other authors (EM, VG) then conducted a thematic analysis guided by the work of Braun and Clarke [20]. We audio recorded interviews and transcribed them verbatim. Using NVivo software (version 12.0), two authors (FP, EM) independently codified the content of each interview and generated initial codes. At first, codes were deductively identified using the NPT constructs. However, not all elements from the interviews fit neatly into the NPT framework, and subsequently the text was then inductively coded to enrich the analysis. The same two authors (FP, EM) used the constant comparative method to discuss their coding and content interpretation. A third author (VG) resolved any disagreements. Cohen’s kappa statistic was calculated to measure inter-rater agreement for coding. When agreement was unsatisfactory (kappa < .7) [21], FP and EM deliberated coding again to reach consensus. As coding progressed, the team iteratively searched for prevalent themes within each construct. The prevalence of a theme was determined according to its frequency within the interview transcripts.

Themes were reviewed and underwent refinement of their definitions throughout the analytic process.

## Results

We interviewed 10 ED physicians (50 % women; 60% certified in Emergency Medicine by the College of Family Physicians of Canada (See Table 1).

**Table 1.**
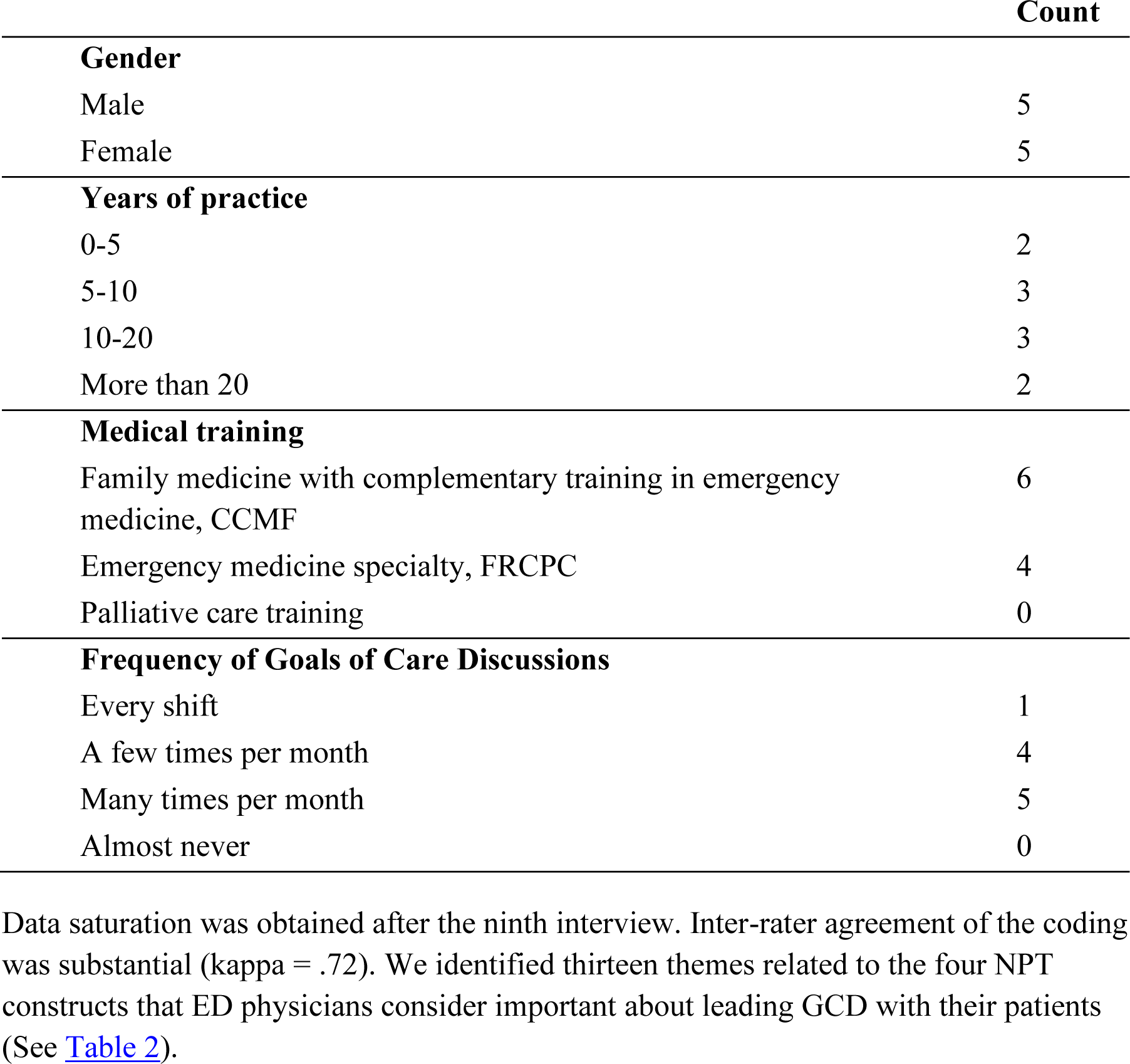
Characteristics of participating physicians.

|  | Count |
| --- | --- |
| **Gender** |  |
| Male | 5 |
| Female | 5 |
| **Years of practice** |  |
| 0-5 | 2 |
| 5-10 | 3 |
| 10-20 | 3 |
| More than 20 | 2 |
| **Medical training** |  |
| Family medicine with complementary training in emergency medicine, CCMF | 6 |
| Emergency medicine specialty, FRCPC | 4 |
| Palliative care training | 0 |
| **Frequency of Goals of Care Discussions** |  |
| Every shift | 1 |
| A few times per month | 4 |
| Many times per month | 5 |
| Almost never | 0 |

### Coherence

We identified six themes related to the construct of coherence (See Table 2 for all constructs, themes, and examples for each).

**Table 2.**
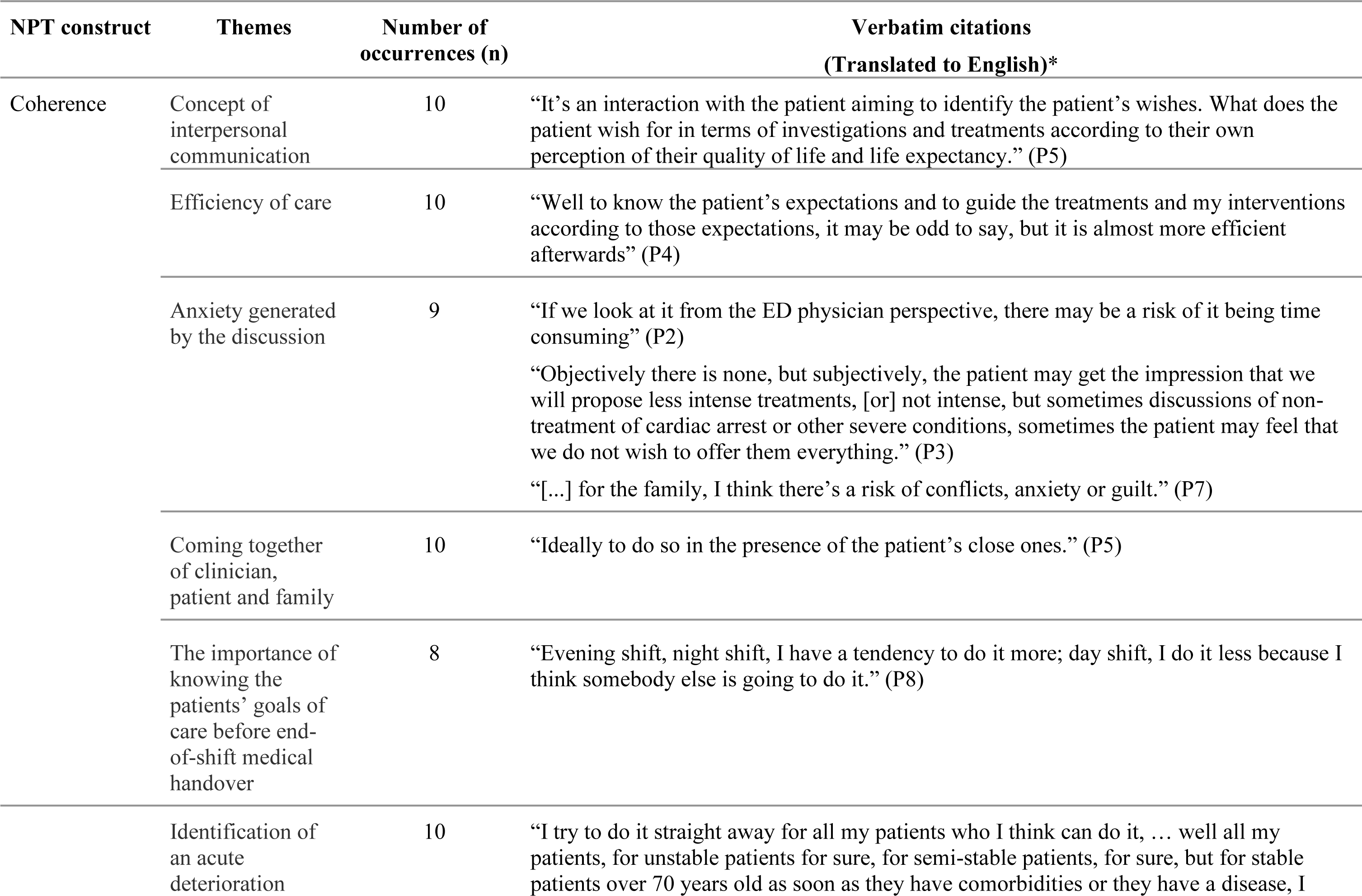
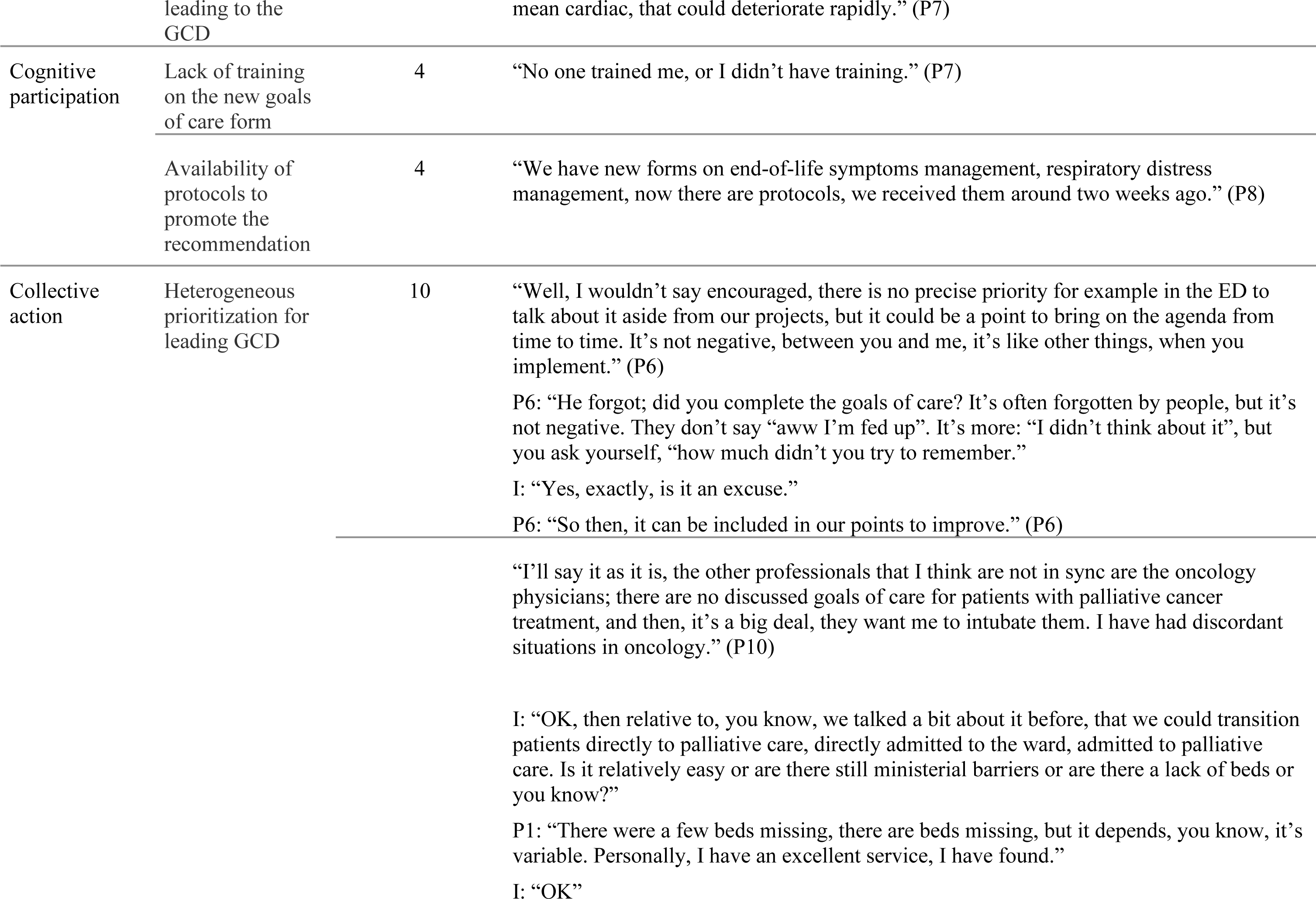
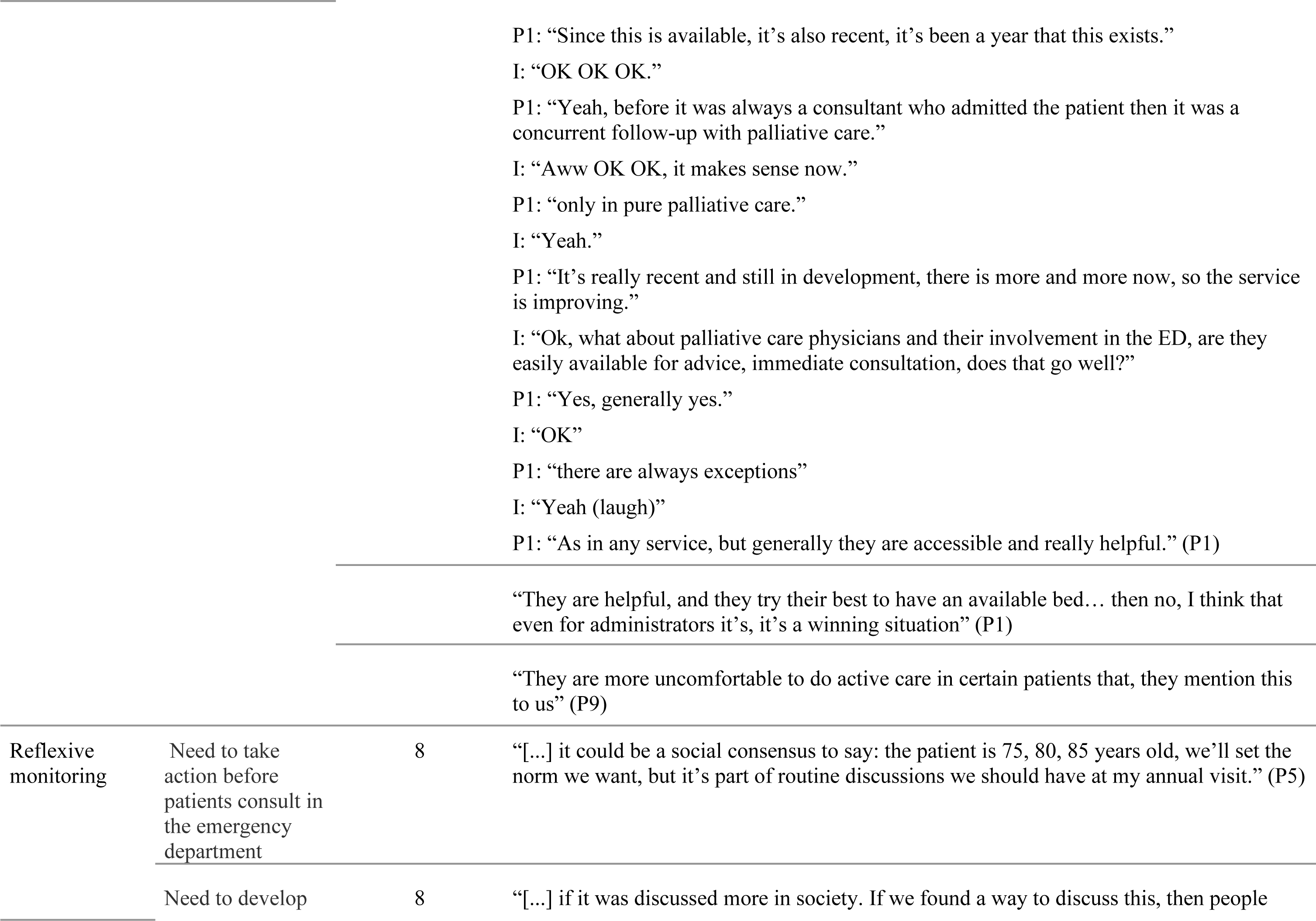
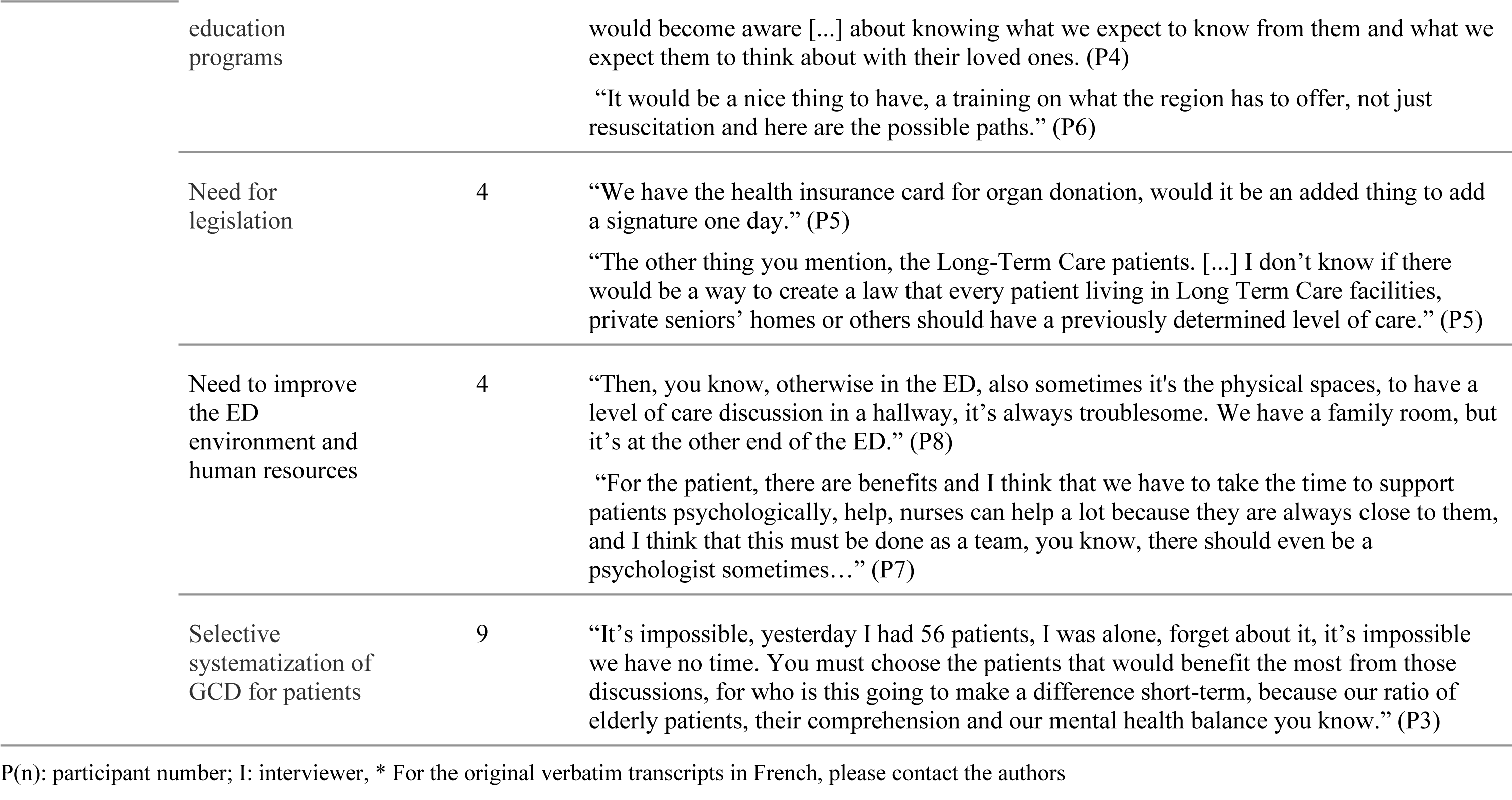
Themes related to the four Normalization Process Theory constructs.

#### Interpersonal communication

All participants (n = 10) defined GCD as the verbal interactions between a physician and a patient or surrogate decision maker capable of discussing the patient’s wishes and expectations about end-of-life care, particularly about the limitations or lack of limitations guiding care decisions about investigations and/or treatments. This discussion allows the patient, or surrogate decision maker and the treating physician to establish a mutual understanding about the desired care in anticipated medical situations. However, most participants (n = 6) indicated facing difficulties explaining the exact difference between the four goals of care on the INESSS form (Appendix 3), which raises the question of whether dividing the goals of care into four categories is helpful or detrimental to the discussion.

Participants explained how they communicated with their patients during GCD. Some (n = 2) approach goals of care, resuscitation, and endotracheal intubation independently, while others include all these discussion topics in the same discussion (n = 1). Most (n = 6) approach the discussion using a directive informed assent approach by suggesting interventions and treatments according to the goal of care they identified as appropriate after reviewing the patient’s comorbidities. Emergency physicians then clarify their reasoning with the patient and specify that the patient’s values and expressed preferences lead to a reconsideration of the suggested GCD.

#### Efficiency of care

GCD was acknowledged by all participants (n = 10) as an important process in the delivery of efficient care. A GCD guides the medical team in defining the orientation of care and ensures that patients’ values, goals, and expectations surrounding end-of-life care are taken into consideration. It also allows patients to express their preferences to their family. Participants also suggested that goals of care discussions lead to optimal resource use and reduction of costs in the health system by preventing the use of non-beneficial treatments.

#### Anxiety generated by the discussion

Physicians’ and patients’ anxiety were identified as a barrier to leading GCD in the ED (n = 9). ED physicians mentioned that the lack of time was a main source of concern. Another important source of concern and moral distress for our participants was when patients expressed unrealistic expectations about their prognosis or the benefit of certain aggressive life-sustaining therapies. According to our participants, patients may also face moral distress when family members express discordant values and preferences concerning the patient’s goals of care. Some physicians (n = 2) also felt that patients may even fear being abandoned if they decide on less aggressive goals of care. Others (n = 4) also expressed concerns that these discussions might induce guilt in family members.

#### Identification of an acute deterioration leading to the GCD

When questioned about how they prioritize which patient would benefit from a GCD, all participants (n = 10) stated that they assess the risk of acute deterioration to guide whether they should engage in a GCD with their patient. Our participants stated that they also use the patient’s characteristics, such as a patient’s age, frailty, and comorbidities to help them decide whether to engage in GCD. For stable patients who will be hospitalized, most ED physicians (n = 6) defer the GCD to the consulting treating team. However, a few participants (n = 4) mentioned engaging in GCD with chronic terminally ill patients who frequently visit the ED and for which treatment options are running out. The participants stated that GCD were not required for stable patients presenting a benign complaint and for patients where maximal care was unequivocal (e.g., young and healthy patients). For patients unable to consent without any available representative, participants make every effort to contact a substitute decision maker, but unfortunately urgent decisions sometimes need to be made for these patients without any guidance from a previous GCD.

#### Meeting between clinicians, patients and family / the importance of knowing the patients’ goals of care before medical handover

According to all our participants (n = 10), two major factors influence the timing to start the discussion of a GCD. The first is when a clinician, a patient and family members meet in the ED and the second is knowing the patient’s goals of care before end-of-shift medical handover. Also, if the patient’s condition is unstable and urgent critical care is needed or if the patient may soon lose their capacity to consent, ED physicians will initiate a GCD as soon as possible.

### Cognitive participation

We identified two themes related to the construct of cognitive participation.

#### Lack of training

Nearly all (n = 9) participants highlighted a lack of training on the standardized INESSS goals of care form (See Appendix 3) implemented in 2016.

#### Availability of protocols

In the case that palliative care is chosen as a patient’s preferred option during the GCD, participants mentioned that an end-of-life respiratory distress care protocol helps standardize care and ensures patient-centered transitions between acute care and hospice in addition to palliative care. Also, one participant mentioned that the electronic order entry system built-in with the ED information system (i.e., Med-Urge, *MédiaMed Technologies*, Mont-Sainte-Hilaire, Québec) allows physicians to indicate and inform all care team members about patients’ preferred goals of care.

### Collective action

We only identified one theme under the construct of collective action.

#### Heterogeneous prioritization for leading GCD

Collectively, engagement towards GCD is a heterogeneous priority among various ED stakeholders. Participants agreed that this intervention is supported and encouraged by the team of ED physicians, but oversight and disengagement are still frequent. According to our ED physician participants, the nursing team is an invaluable ally for GCD. The nursing team to the same degree expresses interest in being included in determining goals of care to allow continuity and congruence of care with the patient’s desires. Some participants (n = 4) mentioned that certain medical specialties (e.g., respirology and cardiology) conduct more GCD with their terminally ill patients than other specialties. For example, some participants (n = 4) pointed to a frequent discrepancy between the palliative nature of certain oncology treatments and the patient’s lack of understanding of the incurable nature of certain terminal cancer conditions. They also criticized the frequent lack of documented GCD with many terminally ill oncology patients. Moreover, participants perceived that hospital administrators were working to facilitate GCD but did not consider GCD as their main priority.

### Reflexive monitoring

We identified five themes under the construct of reflexive monitoring.

#### Need to take action before patients consult in the ED

ED physicians (n = 8) suggested to encourage collaboration in GCD between primary care professionals, ED physicians and other hospital physicians. The objective of which is to promote the use of goals of care in pre-hospital care and to include routine GCD in regular follow-up visits with primary care physicians.

#### Need to develop education programs

Participants (n = 8) mentioned the need to improve the population’s general understanding of GCD, to offer medical training on GCD, end-of-life care and care trajectories, and to standardize the approach to leading GCD.

#### Need for legislation and selective systematization of GCD for patients

ED physicians (n = 4) suggested including documentation about a patient’s goals of care in the provincial *Dossier Santé Québec*, in notarized documents, signed on their health insurance card, and in the patient’s hospital record. In addition, physicians should be required to lead GCD in public and private long-term care facilities, and when caring for high-risk populations such as oncology patients and patients being admitted to hospital.

#### Need to improve the ED environment and human resources

Participants (n = 4) proposed to implement a psychology service in the ED and to promote adapted physical premises for GCD.

#### Selective intervention

To optimize the implementation of GCD, participants (n = 9) suggested defining and systematizing the selection of patients that are the most likely to benefit from GCD.

## Discussion

We aimed to explore emergency physicians’ opinions about the feasibility of leading GCD in their daily practice as suggested by clinical guidelines. Using the Normalization Process Theory (NPT), we identified positive and negative factors influencing the feasibility of leading GCD in the ED.

ED physicians interviewed during this study understand the principles of GCD and recognized that this practice is well-established and should be a part of their standard medical practice. According to the NPT, implementation can be facilitated if healthcare professionals understand the intervention and its importance. Our participants also recognized that care efficiency can be improved when conducting GCD appropriately. ED physicians valued the improved transitions in care and quality of care due to earlier and better GCD in the ED. Above all, early and high quality GCD are beneficial for patients and their family by improving end-of-life care, reducing anxiety, and avoiding unnecessary interventions [5]. A transition towards palliative care when appropriate is also associated with reductions in hospital costs, fewer ED visits, shorter hospitalizations and fewer hospital deaths [8,22,23].

Even if GCD were generally well perceived and that participants were willing to implement this practice, several sources of improvement are required before GCD can be fully and efficiently implemented in the ED. Similar to previous systematic reviews [24] and ED-specific studies [25], our results suggest that GCD can be done in the ED. Similar to other work [25,26], we also identified barriers specific to the ED context that hinder GCD. Anxiety, time constraints, moral distress, absent family members, and lack of training and gaps in information continuity between primary, secondary, and tertiary care all contribute to avoidance of GCD in the ED. A multiprofessional approach may alleviate some of this distress by allowing patients and their families to address unrealistic expectations of prognosis or difficult emotions arising from moral distress [27]. Such professionals could also serve as mediators or neutral third parties in navigating and negotiating terms of GCD with patients.

In their interviews, our participants highlighted the absence of standardized techniques to approach GCD along with variation in the identification of patients requiring GCD as barriers. As such, ED physicians approach the task of GCD relying on their values and preferences, which may contribute to a heterogeneous GCD practice. ED physicians in our study used an informed assent approach in which they limit options presented to patients, like ‘nudging’ patients to make decisions based on ethically acceptable options [28]. Existing resources could also be used to standardize an informed assent approach. Other authors suggest communication models [22] such as the SILVER method [29], the SPIKES model [30], the Serious Illness Conversation Guide [31–33], and the communication course for emergency physicians developed by Grudzen and colleagues [34] as tools to help GCD implementation through effective physician-patient communication and physician’s behaviors while approaching and discussing GCD. The communication aspect of GCD is a determining factor in end-of-life care because it allows for diagnosis, treatment, and prognosis comprehension in addition to the reduction of traumatic stress for family and next of kin [5]. Patient perceptions of cardiovascular resuscitation are often incomplete and as such, physicians in collaboration with broader interprofessional teams [35] must explain the medical implications of those interventions [36] while balancing the uncertainty of a patient’s prognosis [7,25].

Other clinical tools exist to support physicians in their decision to get involved in a GCD, such as the surprise question: “Would you be surprised if this patient died within the next 30 days?” [37] or the PREDICT score helping physicians systematically identify patients who would benefit from GCD [38]. Populational and professional education programs about GCD were also considered to be a potential facilitator for both physicians and allied professionals. For instance, the Education in Palliative and End-of-life Care for Emergency Medicine (EPEC-EM) course and The Improving Palliative care in Emergency Medicine Project offer resources to integrate end-of-life care in daily ED medical practice [39]. Early exposure to the concept of GCD and advance care planning among long-term care residents and their relatives may improve care transitions for long-term care patients to EDs [40].

In addition to using tools to facilitate the standardization of GCD, system-level change must occur to facilitate the implementation of these interventions. Interprofessional collaboration is a prerequisite because GCD cannot be performed with every ED patient. Routine and systematic GCD by primary care providers supported by better information continuity between healthcare teams poses one solution [25]. Engaging and empowering primary care providers to lead GCD can lead to higher quality and earlier GCD, more home-based end-of-life care, and fewer ED visits [27]. Participants voiced their desire for increased support from interprofessional teams and certain specialists, and better access to palliative care teams. Some delegation of responsibilities has occurred: since 2021, nurse practitioners in Québec can complete goals of care with their patients [41].

An increased access to adequate and timely GCD will necessitate collective social action initiatives. Although the Québec government instituted the Act Respecting End-of-Life Care in 2015 [42], some participants called for additional legal reforms that would mandate that organizations like long-term care institutions bear some responsibility to document GCD for their patients. Since the COVID-19 pandemic, governmental initiatives have increased the support for more complete and earlier GCD before patients arrive in the ED and face potentially difficult decisions [41].

In February 2024, the INESSS updated their recommendations about GCD [11]. Some recommendations propose changes relevant to the points raised by our participants. First is the simplification of the form to three non-hierarchical categories (i.e., Prolongation (P), Equilibrium (E), and Comfort (C)) instead of four hierarchical levels of care (i.e., A (Prolong life with all necessary care), B (Prolong life with some limitations to care), C (Ensure comfort as a priority over prolonging life), and D (Ensure comfort without prolonging life)). While Prolongation and Comfort are distinct, there is a considerable gray area when considering Equilibrium. INESSS recommends that information pertinent to choosing this goal of care should be documented clearly and in advance to guide the care team in an emergency situation. INESSS has also recommended the initiation of GCD throughout care, especially as soon as a serious chronic diagnosis is identified, but also including high-risk acute situations and hospital admissions, in alignment with the perceptions of our participating ED physicians. The INESSS recommendations currently do not provide a detailed overview of practices to identify specific field issues, recommend actions, nor propose monitoring indicators.

ED physicians in our study positively perceived GCD, despite identifying important constraints in practice. Future studies will be needed to investigate how other ED healthcare professionals such as nurses and social workers [31] can support earlier and better GCD in the ED. Continuous monitoring of the frequency and quality of GCD in the ED will be essential to support ongoing quality improvement and sustainable change in these interventions.

### Limitations and Strengths

Our study has limitations. This study is monocentric, and thereby influenced by the specific characteristics of the local culture and community. Using a convenience sample of motivated ED physicians engaged in improving goals of care discussions in the ED may indicate a selection bias. Their interest and dedication allowed us to collect rich insights in improving GCD.

This study also has strengths. We used a well-documented and tested methodological approach [12] to analyze a complex and prevalent implementation problem across Canada [25,43]. Our results accord with current national and international scientific literature. Using the NPT may allow other investigators to use our results and compare them to diverse cultural, organizational, and legal contexts. Our qualitative analysis of a complex intervention allowed us to expand on solutions with participating physicians to improve the implementation and sustainability of high-quality goals of care discussions in the ED.

### Clinical implications

This study identified facilitating factors for implementation of GCD in the everyday practice for ED physicians. Physicians suggested educating patients on the concept of GCD, imposing legislation on the selective systematization of GCD for patients, and ideally delegating the documentation of patients’ preferences to primary care before any visit to the ED. Other suggestions included training ED physicians on efficient use of the tool, encompassing professional support and institutional prioritization in practicing these discussions. Considering our results, more institutional and provincial support is needed to implement immediate actions aiming to normalize GCD in the ED and pre-ED.

### Research implications

Emergency physicians believe that it is essential to clarify goals of care with specific ED patients. Easy to access quality indicators are needed to document the quality and monitor the frequency of GCD in the ED.

### Conclusion

Goals of care discussions are possible and essential with selected patients in the ED. Nevertheless, clinician-level, collective and policy-making efforts remain essential to ensure the implementation of clinical guideline recommendations.

## Data Availability

Anonymized data produced in the present study are available upon reasonable request to the authors. Please note that interview transcripts were recorded and coded in French.

## Acknowledgements

We thank all participants who generously accepted to participate in this study.

## Authors’ contributions

FP and PA obtained funding for the study from the Department of Family and Emergency Medicine, Université Laval. This study was also supported by an Embedded Clinician Salary Award (ECRA) awarded to PA from the Canadian Institutes for Health Research (CIHR) (#201603) and a Fonds de recherche du Québec – Santé (FRQS) Senior Clinical Scholar Award (#283211). FP, AP and PA designed the study. FP, EM and VG collected and interpreted the data. FP and VG wrote the paper with input from all authors. AN reviewed the manuscript and translated the verbatims from French to English. ML and NG critically reviewed the manuscript and provided improvements to the manuscript. The study findings and final manuscript were critically revised and approved by all authors. The study findings and final manuscript were critically revised and approved by all authors.

## Appendix 1: Standards for Reporting Qualitative Research (SRQR)*

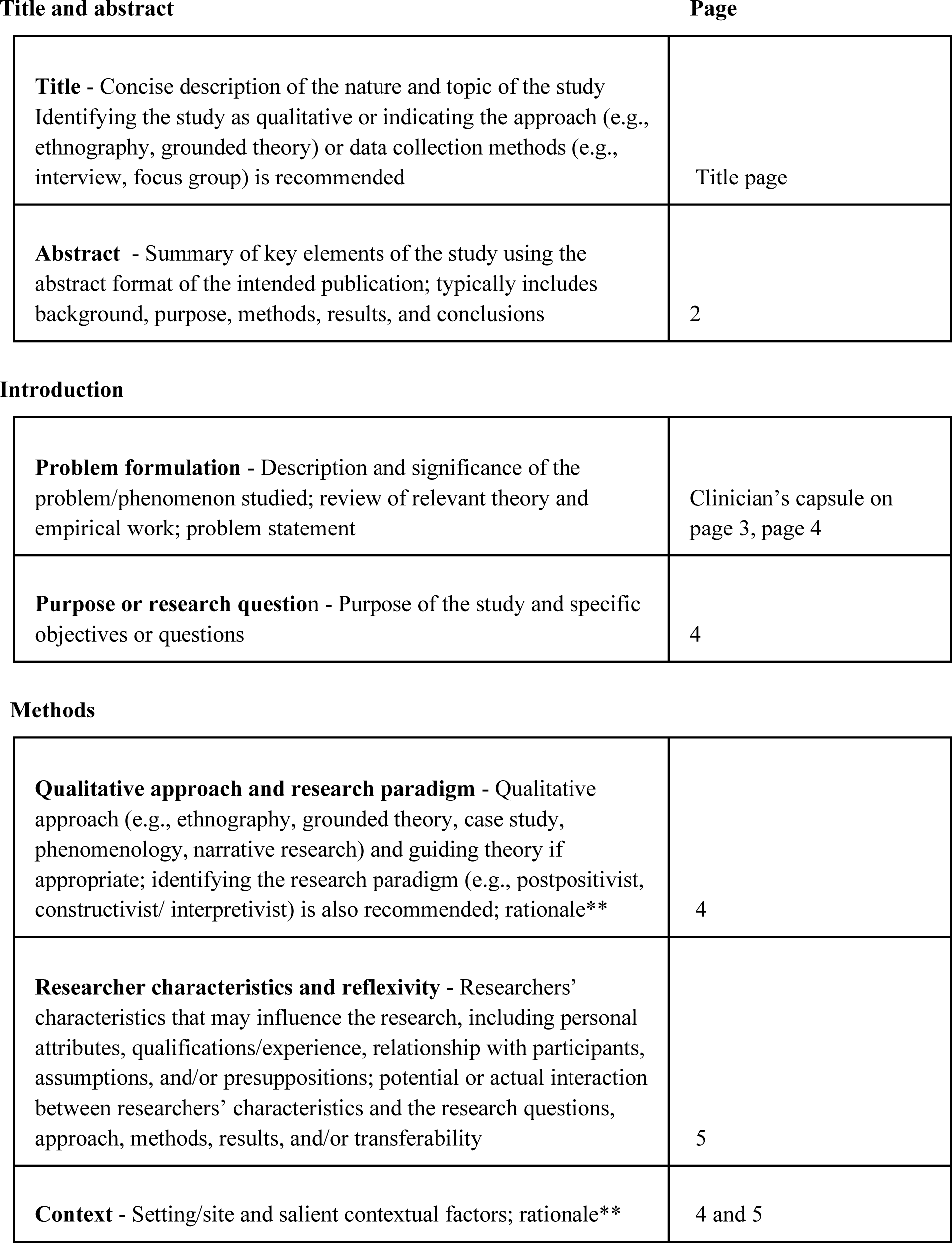
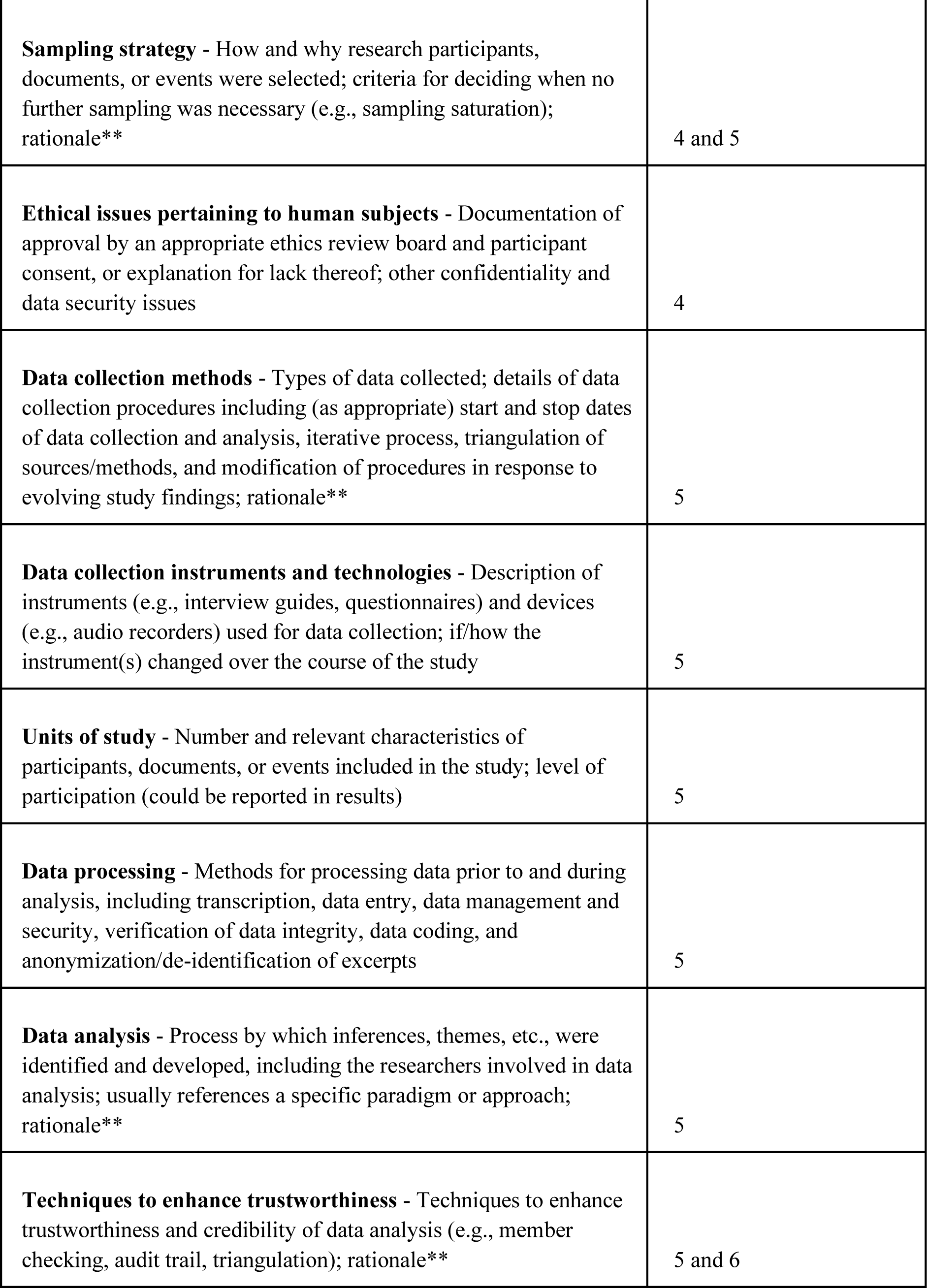
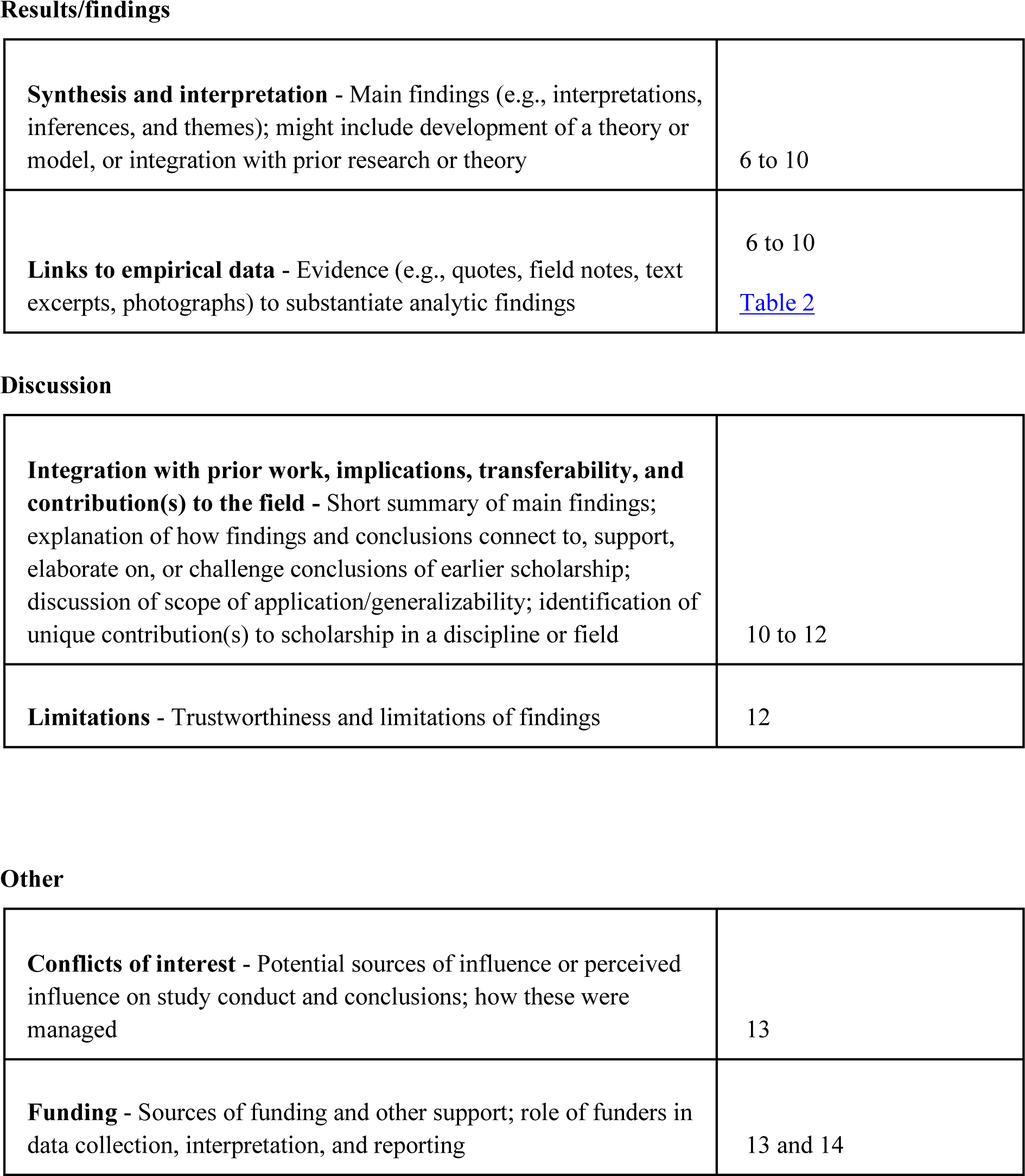
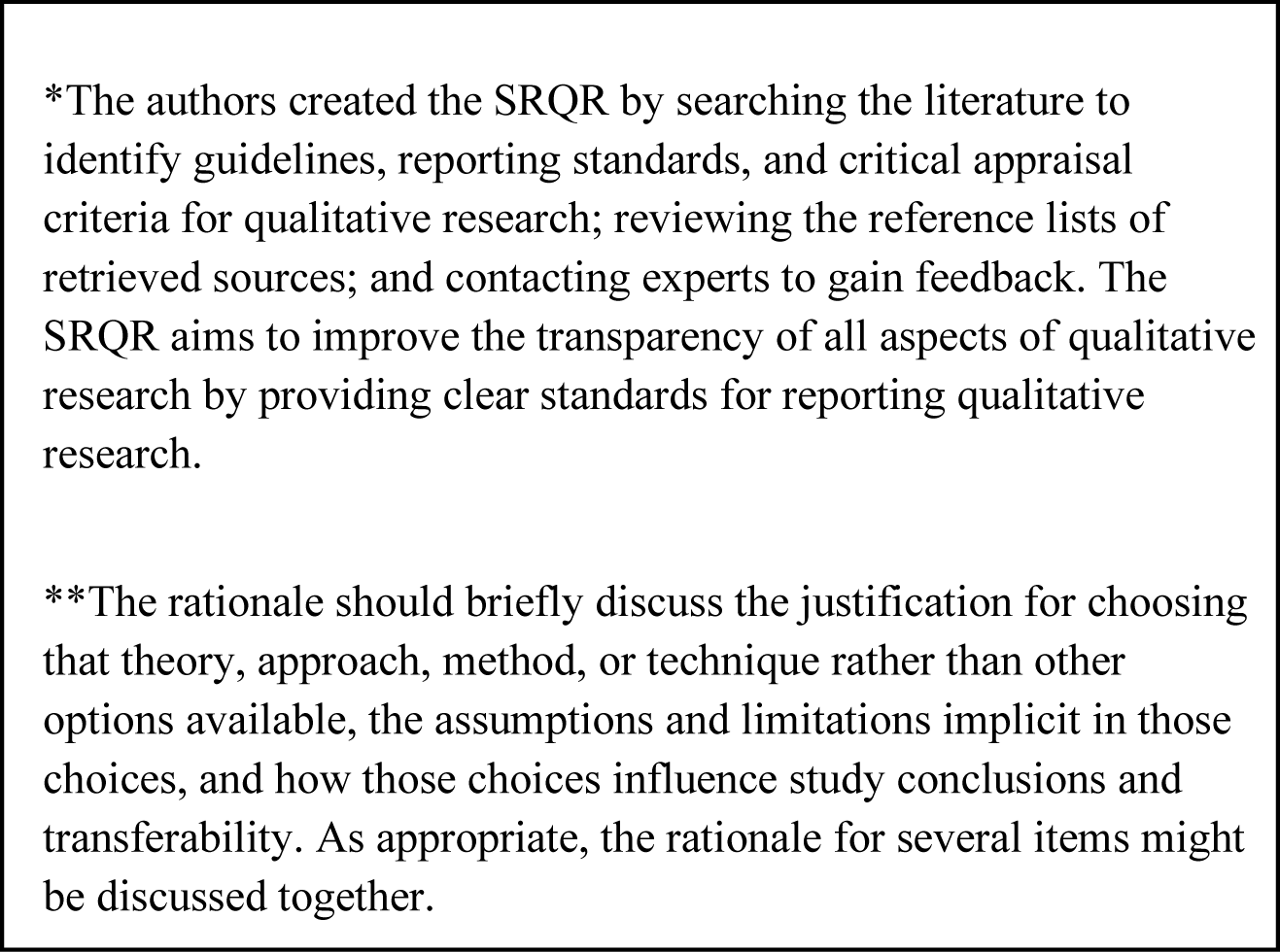

Page
|  |  |
| --- | --- |
| **Title** - Concise description of the nature and topic of the study<br>Identifying the study as qualitative or indicating the approach (e.g., ethnography, grounded theory) or data collection methods (e.g., interview, focus group) is recommended | Title page |
| **Abstract** - Summary of key elements of the study using the abstract format of the intended publication; typically includes background, purpose, methods, results, and conclusions | 2 |
| Introduction |  |
| **Problem formulation** - Description and significance of the problem/phenomenon studied; review of relevant theory and empirical work; problem statement | Clinician's capsule on page 3, page 4 |
| **Purpose or research question** - Purpose of the study and specific objectives or questions | 4 |
| **Qualitative approach and research paradigm** - Qualitative approach (e.g., ethnography, grounded theory, case study, phenomenology, narrative research) and guiding theory if appropriate; identifying the research paradigm (e.g., postpositivist, constructivist/ interpretivist) is also recommended; rationale** | 4 |
| **Researcher characteristics and reflexivity** - Researchers' characteristics that may influence the research, including personal attributes, qualifications/experience, relationship with participants, assumptions, and/or presuppositions; potential or actual interaction between researchers' characteristics and the research questions, approach, methods, results, and/or transferability | 5 |
| **Context** - Setting/site and salient contextual factors; rationale** | 4 and 5 |
| **Sampling strategy** - How and why research participants, documents, or events were selected; criteria for deciding when no further sampling was necessary (e.g., sampling saturation); rationale** | 4 and 5 |
| **Ethical issues pertaining to human subjects** - Documentation of approval by an appropriate ethics review board and participant consent, or explanation for lack thereof; other confidentiality and data security issues | 4 |
| **Data collection methods** - Types of data collected; details of data collection procedures including (as appropriate) start and stop dates of data collection and analysis, iterative process, triangulation of sources/methods, and modification of procedures in response to evolving study findings; rationale** | 5 |
| **Data collection instruments and technologies** - Description of instruments (e.g., interview guides, questionnaires) and devices (e.g., audio recorders) used for data collection; if/how the instrument(s) changed over the course of the study | 5 |
| **Units of study** - Number and relevant characteristics of participants, documents, or events included in the study; level of participation (could be reported in results) | 5 |
| **Data processing** - Methods for processing data prior to and during analysis, including transcription, data entry, data management and security, verification of data integrity, data coding, and anonymization/de-identification of excerpts | 5 |
| **Data analysis** - Process by which inferences, themes, etc., were identified and developed, including the researchers involved in data analysis; usually references a specific paradigm or approach; rationale** | 5 |
| **Techniques to enhance trustworthiness** - Techniques to enhance trustworthiness and credibility of data analysis (e.g., member checking, audit trail, triangulation); rationale** | 5 and 6 |
| Results/findings |  |
| **Synthesis and interpretation** - Main findings (e.g., interpretations, inferences, and themes); might include development of a theory or model, or integration with prior research or theory | 6 to 10 |
| **Links to empirical data** - Evidence (e.g., quotes, field notes, text excerpts, photographs) to substantiate analytic findings | 6 to 10<br>Table 2 |
| **Integration with prior work, implications, transferability, and contribution(s) to the field** - Short summary of main findings; explanation of how findings and conclusions connect to, support, elaborate on, or challenge conclusions of earlier scholarship; discussion of scope of application/generalizability; identification of unique contribution(s) to scholarship in a discipline or field | 10 to 12 |
| **Limitations** - Trustworthiness and limitations of findings | 12 |
| Other |  |
| **Conflicts of interest** - Potential sources of influence or perceived influence on study conduct and conclusions; how these were managed | 13 |
| **Funding** - Sources of funding and other support; role of funders in data collection, interpretation, and reporting | 13 and 14 |

\*The authors created the SRQR by searching the literature to identify guidelines, reporting standards, and critical appraisal criteria for qualitative research; reviewing the reference lists of retrieved sources; and contacting experts to gain feedback. The SRQR aims to improve the transparency of all aspects of qualitative research by providing clear standards for reporting qualitative research.
\*\*The rationale should briefly discuss the justification for choosing that theory, approach, method, or technique rather than other options available, the assumptions and limitations implicit in those choices, and how those choices influence study conclusions and transferability. As appropriate, the rationale for several items might be discussed together.

## Appendix 2: Interview guide (English translation)*

In January 2016, INESSS made recommendations in its report of levels of care: quality norms and standards, or *Les niveaux de soins : cadre, processus et méthodes d’élaboration du guide sur les normes et standards de qualité*. In general, a discussion of levels of care should be initiated with anyone whose current prognosis suggests a lack of improvement or a lasting deterioration in their health, quality of life, or autonomy, in the short or medium term. This recommendation is particularly targeted for people who:

1. have a limited life expectancy
2. are at a high risk of deterioration due to an acute or chronic health condition or a state of frailty
3. at risk of major complications from surgery, endoscopy or other invasive procedures
4. are receiving palliative or end-of-life care
5. are struggling with proven cognitive impairment
6. are followed in oncology and who have a limited prognosis
7. have a high probability of being hospitalized or admitted at the emergency department within a year
8. are admitted at the emergency room, acute care or intensive care
9. have completed and registered an advance medical directive (AMD)
10. request a discussion of levels of care.

The following interview is intended to understand whether this recommendation is feasible.

## Before starting the interview

1. Discuss the length of the interview (approximately 30 minutes) and content of the interview
2. Ask for permission to record
3. Informed consent was already obtained as part of a larger study

*Interview questions organized by the NPT constructs (coherence, cognitive participation, collective action, and reflexive monitoring)*.

## Coherence

1. What do you think a levels of care discussion is?
2. In your opinion, what are the risks and benefits of discussing levels of care with your patients? *Sub-questions: (patient, professional, systems, population)*.
3. At what point during the ICU stay (or emergency department, or other place of care) do you determine *<u>that it is necessary</u>* to initiate a level of care discussion with a patient?
4. At what point during the ICU stay (or emergency department, or other place of care) do you determine *<u>that it is not necessary</u>* to initiate a level of care discussion with a patient?
5. In what medical situations do you think these discussions *<u>should be</u>* undertaken at the outset?
6. In what medical situations do you think these discussions *<u>should be avoided</u>* in the first place?

## Cognition

7. What has been put in place to help you implement this recommendation?

## Collective action

8. Do you feel that other health professionals support the application of this recommendation?

### Sub-questions: (nurse, other doctors, manager)?

## Reflexive monitoring

In your opinion, could the practice of level of care discussions be sustained or systematized under current conditions?
What policy or culture changes are needed to facilitate the implementation of the recommendation?

*For the Original French version of this interview guide, the « *Grille d’entrevue semi-dirigée basée sur la Théorie du processus de normalisation* », please contact the authors.

## Appendix 3: *Institut national d’excellence en santé et services sociaux de Québec* form: Levels of care and cardiopulmonary resuscitation (English)*

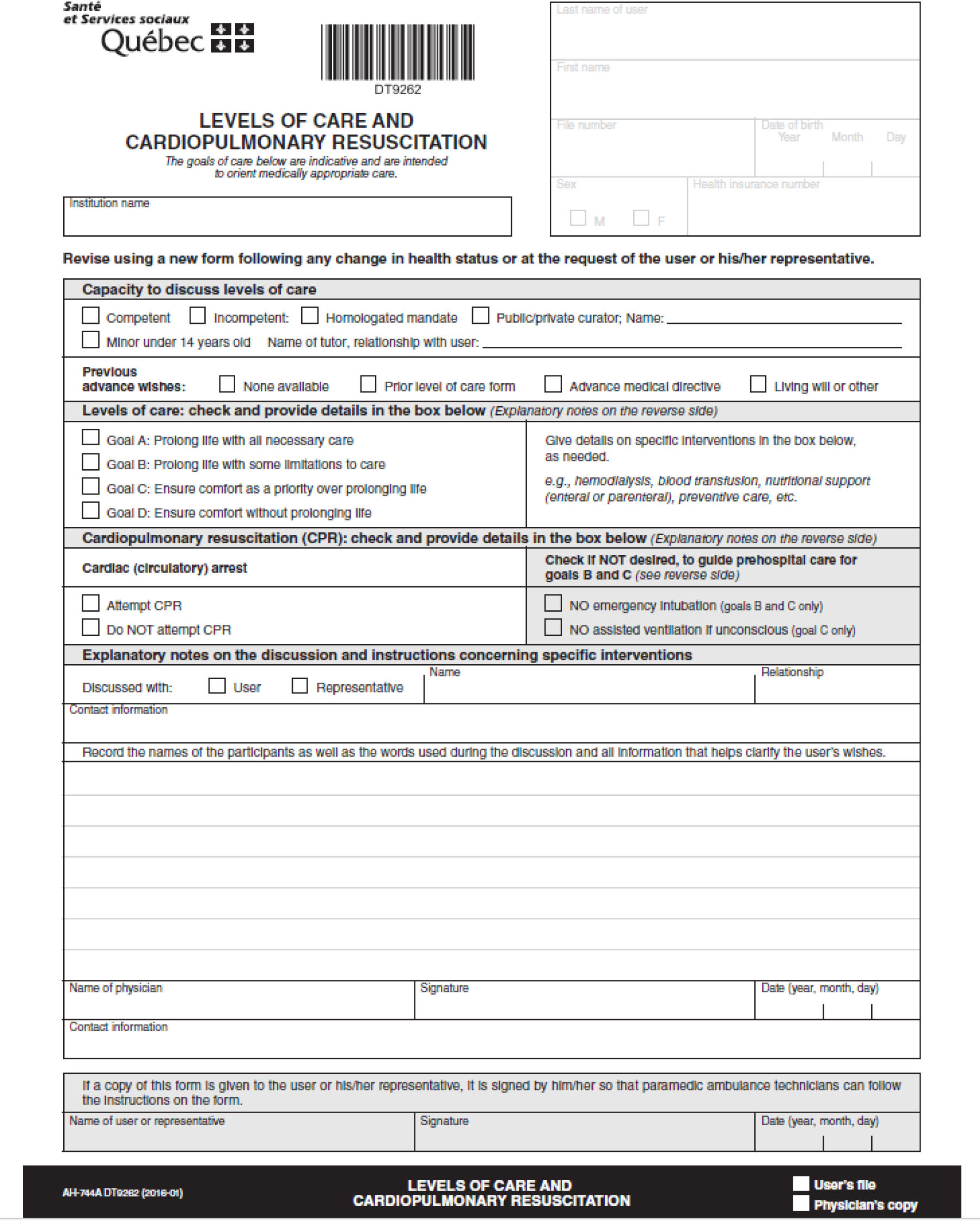
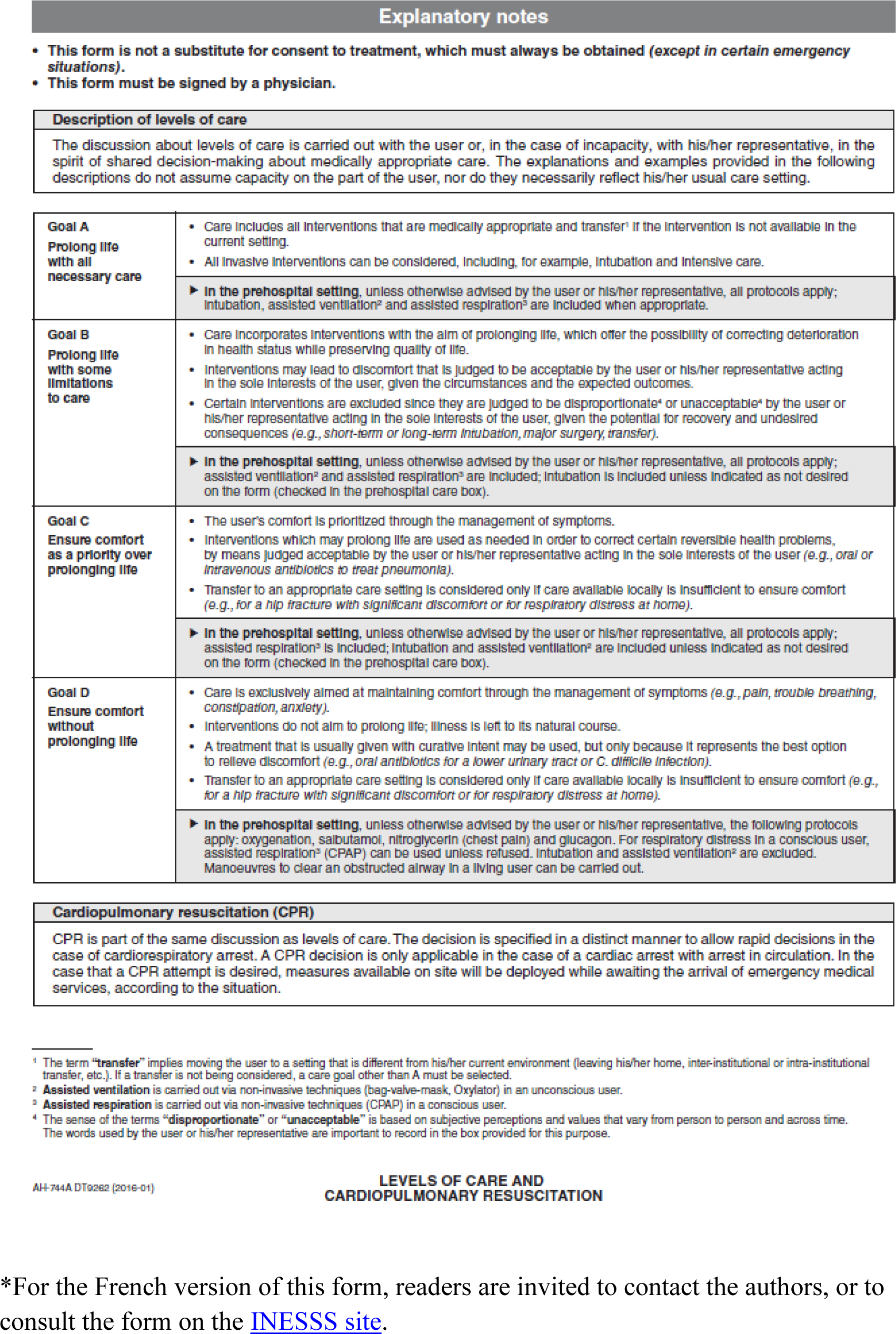

